# SARS-CoV-2 serosurvey in Health Care Workers of the Veneto Region

**DOI:** 10.1101/2020.07.23.20160457

**Authors:** Plebani Mario, Padoan Andrea, Fedeli Ugo, Schievano Elena, Vecchiato Elena, Lippi Giuseppe, Lo Cascio Giuliana, Porru Stefano, Palù Giorgio

## Abstract

**Background:** The ongoing outbreak of coronavirus disease (COVID-19) caused by the severe acute respiratory syndrome coronavirus 2 (SARS-CoV-2) poses formidable challenges to all health care systems. Serological assays may improve disease management when appropriately used, for better understanding the antibody responses mounted upon SARS-CoV-2 infection and for assessing its real prevalence. Although testing the whole population is impratical, well-designed serosurveys in selected subpopulations in specific risk groups may provide valuable information.

**Aim:** we evaluated the prevalence of SARS-CoV-2 infection in health care workers who underwent molecular testing with reverse transcription real-time polymerase chain reaction (rRT-PCR) in the main hospitals of the Veneto Region by measuring specific antibodies (Abs).

**Methods:** both IgM and IgG antibodies against SARS-Cov-2 S-antigen and N-protein were measured using a validated chemiluminescent analytical system (CLIA) called Maglumi™ 2000 Plus (New Industries Biomedical EngineeringCo., Ltd [Snibe], Shenzhen, China)

**Results:** A total of 8285 health care workers were tested. SARS-CoV-2 specific antibodies (IgM, IgG or both) were detectable in 378 cases (4.6%, 95% CI 4.1-5.0%). Seroconversion was observed in 4.4% women and 5% men, but the difference was not significant. Although detectable antibodies were found in all severe COVID-19 patients (100%), lower seropositivity was found in mild disease (83%) and the lowest prevalence (58%) was observed in asymptomatic subjects.

**Conclusion:** Seroprevalence surveys are of utmost importance for understanding the rate of population that has already developed antibodies against SARS-CoV-2. The present study has the statistical power to define precisely the circulation of SARS-CoV-2 in a cohort of health workers in our region, with its prevalence (4.6%) reflecting a relatively low circulation. Symptomatic individuals or those hospitalized for medical care were 100% antibody positive, whilst Abs were only detectable in 58% of asymptomatic carriers.

## Introduction

The spread of coronavirus disease 2019 (COVID-19), caused by severe acute respiratory syndrome coronavirus 2 (SARS-CoV-2), has become a pandemic, with sustained human-to-human transmission. Since the initial identification of COVID-19 in December 2019, there has been an exponential rise in the number of cases worldwide, especially in Italy (1). The reasons for the rapid spread include the high transmissibility of the virus, especially among asymptomatic or minimally symptomatic carriers, as well as the apparent absence of cross-protective immunity from related coronavirus infections, and the tardy public health response measures (2, 3). Nucleic acid tests for detecting SARS-CoV-2 RNA genome are widely employed for diagnosing COVID-19 (4), but cannot be deployed for prevalence investigations or for sensing prior exposure to an infectious agent. Therefore, there remains a great need for assays that measure antibody responses and determine seroconversion (5). In fact, inadequate knowledge about the extent of COVID-19 pandemic challenges public health response and planning. Although it is impractical to test the entire population, well-designed serosurveys play a relevant role to determine how prevalent COVID-19 may be in the general population, especially in selected high risk groups (e.g. health care workers) (6, 7). The several serological assays developed and currently available on the market differ for two major variables: a) the format used and, in particular, whether the test is a quantitative laboratory-based immunoassay ELISA, CLIA or a qualitative point-of-care test (POCT); and b) the SARS-CoV-2 antigen targeted, in particular if the antibodies are addressed against the spike surface protein (s) (namely subunit 1 and 2), and/or the spike receptor binding domain (RBD), and/or the nucleocapside protein (N) (6). In addition, they may detect total antibodies (tAb), and/or one or more specific immunoglobulins such as IgA, IgM, and IgG according to the main purposes of the measurements. Therefore, this study was aimed to determine the prevalence of COVID-19 in the health care workers of the Veneto Region through the measurement of IgM and IgG SARS-CoV-2 antibodies using a validated chemiluminescent assay (CLIA) (8).

## Materials and methods

### Subjects included in the study

This multicenter study involved several Structures of the National Healthcare Service of the Veneto Region, in which was reported an early cluster of SARS-CoV-2 infection in Italy on February 21th, 2020. In particular, serological determinations were performed at the University Hospital of Padova, University Hospital of Verona and S. Bortolo Hospital of Vicenza laboratories. A total of 8285 healthcare workers were included in the study. For each patient a serum sample was collected, heat inactivated at 56°C for 30 minutes before the analyses. To evaluate sensitivity molecular rRT-PCR tests performed since February 22 to 13 days before serological determinations have been considered for all subjects. This interval of 13 days was excluded from the analyses on the basis of the current knowledge of seroconversion time and antibody kinetics (8-10). For estimating specificity, a subject was considered SARS-CoV-2 negative if at least three sequential negative results rRT-PCR were found (one of that within 12 before serological testing). The association between sensitivity of serological test and severity of SARS-Cov-2 infection was assessed through linkage with a regional centralized surveillance system including all infection cases and related hospitalizations. This study was approved by the Regional Committee for Bioethics (prot. n. 178357) and the Ethics Committee of the University Hospital of Verona (prot. n. 22851) and all subjects recruited to the study expressed an informed consent for being enrolled.

### Measurement of SARS-CoV-2 antibodies

Maglumi™ 2000 Plus (New Industries Biomedical EngineeringCo., Ltd [Snibe], Shenzhen, China) is a chemiluminescent analytical system (CLIA) for the detection of both IgM and IgG antibodies against SARS-Cov-2 S-antigen and N-protein. According to the manufacturer’s inserts (271 2019-nCoV IgM, V2.0, 2020-03 and 272 2019-nCoV IgG, V1.2, 2020-02), the clinical sensitivities of IgM and IgG were 78.65% and 91.21%, respectively, while the specificities of IgM and IgG were 97.50% and 97.3%, respectively. Manufacturers’ reported thresholds were 1.0 kAU/L for IgM and 1.1 kAU/L for IgG, respectively. The analytical performance of this system has been evaluated and described elsewhere (8).

## Results

A total of 8285 health care workers from the University-Hospital of Padova and Verona, from the Istituto Oncologico Veneto, from the Hospital of Vicenza and from other healthcare structures have been investigated; 5933 (71.6%) were women and 2352 were men (28.4%). The mean age was 43.2±11.6 yrs, 43.6±11.4 years for women and 42.2±12.5 yars for men, respectively (t=5.06, p<0.001).

SARS-CoV-2 specific antibodies (IgM, IgG or both) were detectable in 378 healthcare workers (4.6%, 95% CI: 4.1-5.0%). In particular, IgM antibodies have been detected in 82 subjects, while IgG have been found in 343 health care workers. Seroconversion was observed in 4.4% women and 5% men, but the difference did not achieve statistical significance (χ^2^=1.41, p=0.493). Table 1 shows the distribution according to different age groups (χ^2^=18.82, p = 0.001). Table 2 shows the seroprevalence distribution according to the different health care role (physicians, nurses and health care assistants, χ^2^=10.55, p = 0.014). A significant higher seroprevalence could be observed in health care assistants compared to other groups (χ^2^=5.34, p = 0.021).

**Table 1:** Total number and percentages of positive tests with 95% confidence intervals (CI), subdivided by age classes.

| Age classes (yrs) | Total number of tests | Percentage (%) of positive tests | Percentage 95% CI |
| --- | --- | --- | --- |
| < 30 yrs | 1512 | 4.1% | 3.2-5.2% |
| 30-39 yrs | 1826 | 3.5% | 2.7%-4.4% |
| 40-49 yrs | 1962 | 4.4% | 3.6%-5.4% |
| 50-59 yrs | 2389 | 6.0% | 5.1%-7.1% |
| > 60 yrs | 596 | 3.7% | 2.3-5.5% |

**Table 2:** Total number and percentages of positive tests with 95% confidence intervals (CI), subdivided by the different health care figures

| Healthcare figures | Total number of tests | Percentage (%) of positive tests | Percentage 95% CI |
| --- | --- | --- | --- |
| Physicians | 2337 | 3.6% | 2.8%-4.4% |
| Nurses | 3230 | 4.7% | 4.0-5.5% |
| Healthcare assistants | 1040 | 6.0% | 4.6%-7.6% |
| Others | 1678 | 4.8% | 3.8%5.9% |

To evaluate the diagnostic specificity, 3110 health care operators with at least three nasopharyngeal swabs negative results, were considered: 3049 resulted negative thus leading an estimated specificity of 98% (CI 97.5-98.5%). Figure 1 shows the prevalence of seropositivity in three different groups of subjects positive to nasopharyngeal swabs according to disease severity (group A hospitalized with severe disease; group B Mild disease; group C asymptomatic subjects). While detectable antibodies were found in all severe COVID-19 patients (100%), a lower seropositivity was found in mild disease (83%) and the lowest prevalence (58%) was observed in asymptomatic subjects. The differences were found to be significant with the Cuzick test (z=4.31, p<0.001). An intermediate seropositivity (66%) was found in quarantined at home subjects with “undefined mild or asymptomatic” disease.

**Figure 1:**
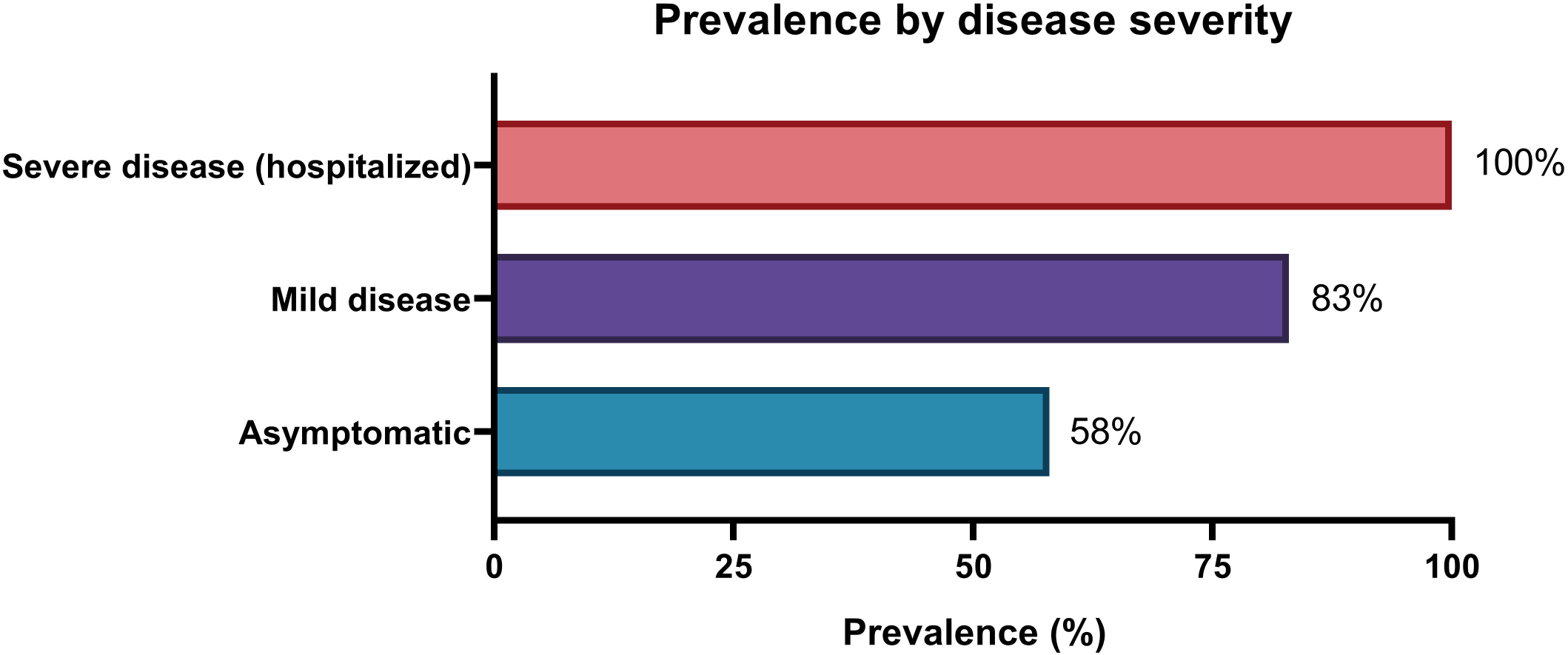
Prevalence of seropositivity in three different groups of subjects, according to disease severity.

## Discussion

Serological testing are essential tools in the management of infectious diseases, including diagnosis of infection, measurement of protective antibody titers upon vaccination, and seroprevalence assessments of immunity in a population. In the case of COVID-19, many issues are still unclear. It is unknown, for example, whether the presence of binding antibody correlates with virus neutralization, whether antibody titers correlate with protection against reinfection and if there is a difference in antibody responses in individuals presenting with severe, mild and asymptomatic COVID-19 (6). While we have to better understand and clarify all these issues, the role of serological testing in assuring information on prevalence of SARS-CoV-2 infection in different populations is unquestionable (11). The present study is a large seroprevalence investigation dealing with exposure to SARS-CoV-2 of personnel attending hospitalized and critically ill patients suffering from COVID-19. The survey was carried out with a CLIA assay endowed with high analytical performances (8-13) and capable to measure (semi)-quantitatively IgM and IgG that recognize both the N and S proteins.

Although the study population cannot be considered representative of the whole healthcare workforce in the Veneto Region, results indicate that, on average, 4,6% of this multicenter cohort seroconverted by production of IgM and IgG. IgGs were present more often than IgMs, a result that is in keeping with a number of other reports (8-15), thus indicating that this isotype is the predominant in term of anti-SARS-CoV-2 immune response.

The prevalence observed in such at risk population reflects the relatively low circulation of SARS-CoV-2 in Veneto Region, much lower compared to other Italian areas where the virus has more widely spread (16). This reflects the overall incidence data of SARS-CoV-2 in Veneto Region, with 20.000 cases out of 5 million inhabitants. On these grounds one could reasonably argue that a minority of general population has been exposed to the virus (2-3%). As a consequence, more than 90% residents are susceptible to viral infection, an estimate that instructs future preventive strategies in case of a new wave of pandemic.

The specificity of the assay was as high as 98%, as assessed in more than 3,000 healthcare workers repeatedly negative at rRT-PCR. A subset of 286 rRT-PCR positive health care professionals, whose blood could be sampled at least 13 days after the nasopharyngeal swab, offered the possibility to test sensitivity of the antibody assay. Only 210 out of 286 blood samples contained detectable anti SARS-CoV-2 IgG and IgM, a result yielding a overall sensitivity of 73%.

Noteworthy, however, a fraction of these health care workers failing to produce antibodies were asymptomatic or with mild symptoms of COVID-19, i.e. 42% and 17%, respectively. Symptomatic individuals or those admitted to hospital for medical care scored 100% antibody positive. This finding confirms previous literature data (17, 18) that indicate a clear correlation between burden of symptoms or disease phenotype and capacity to mount an adaptive immune response. This feature is not unique of COVID-19, but is common to many viral syndromes. In fact, there are always subjects that do not respond to a viral infection with antibody production, but fail to show signs of disease. If this is due to a low sensitivity of the available serological assays or to an individual genetic background is a matter of further investigation that also deals with the nature and strength of innate and adaptive immune responses.

Recent data seem to indicate a relevant role of CD4 and CD8 T cell both in helping production of antibodies against coronaviruses and clearing viral infection from involved tissues (19, 20). The study of cell mediated immunity combined with serology could hence be helpful in predicting COVID-19evolution.

The present study has some limitations. First, the study population cannot be considered representative of the whole healthcare workforce in the Veneto Region, and a selection bias should be necessarily recognized. Second, the relationships of currently measured antibodies with neutralizing activity against SARS-CoV-2 were not evaluated. A body of evidence, however, demonstrates that antibodies targeting different domains of S protein, including S1, RBD and S2, may all contribute to virus neutralization (21, 22). Finally, follow-up data on subjects with detectable SARS-CoV-2 antibodies are not yet available as this follow-up requires some more months to better understand the duration of immune response and protection from reinfection.

## Conclusions

The present study has the statistical power to define precisely the circulation of SARS-CoV-2 in a cohort of health workers. These data, and in particular seroprevalence in our setting, may allow to calculate the predictive positive and negative values, thus overcoming the limitations of diagnostic sensitivity and specificity in clinical practice (23). In addition, our results carry relevant implications for public health policy and control of virus spread in a Region that was badly invested by COVID-19. Moreover, the availability of an assay such as that described in this study can help defining the immune status of an infected host once its values have been normalized to titers obtained by a classical neutralization assay. Quantitative serological assays will be also extremely helpful in better understanding the immune response to SARS-CoV-2 and in assisting development of new vaccines and further developments of new diagnostic and therapeutic tools.

## Data Availability

All data of the seroconversion survey have been reported

## References

1. Onder G, Rezza G, Brusaferro S. Case-Fatality Rate and Characteristics of Patients Dying in Relation to COVID-19 in Italy. JAMA. 2020. (ahead of print); doi:10.1001/jama.2020.4683.

2. Cheng MP, Papenburg J, Desjardins M, Kanjilal S, Quach C, Libman M, et al. Diagnostic testing for Severe Acute Respiratory Syndrome-Related Coronavirus-2: A Narrative Review. Ann Intern Med 2020 (ahead of print); doi:10.7326/M20-1301.

3. Mizumoto K, Kagaya K, Zarebsky A, Chowell G. Estimating the asymptomatic proportion of coronavirus disease 2019 (COVID-19) cases on board the Diamond Princess cruise ship, Yokohama, Japan, 2020. Euro Surveill 2020;25:10.

4. Corman VM, Landt O, Kaiser M, Molenkamp R, Meijer A, Chu DKW, et al. Detection of 2019 novel coronavirus (2019-nCoV) by real-time RT-PCR. Euro Surveill 2020; (ahead of print). doi:10.2807/1560-7917.ES.2020.25.3.2000045

5. Amanat F, Stadlbauer D, Strohmeier S, Nguyen THO, Chromikova V, McMahon M, et al. A serological assay to detect SARS-CoV-2 seroconversion in humans. Nat Med. 2020; (ahead of print). doi:10.1038/s41591-020-0913-5.

6. Krammer F, Simon V. Serology assays to manage COVID-19 Science 2020; (ahead of print). doi:10.1126/science.abc1227. Online ahead of print.

7. Torres R, Rinder HM. Are SARS-CoV-2 serological tests safe right now? Am J Clin Pathol 2020; 153: 709–11.

8. Padoan A, Cosma C, Sciacovelli L, Faggian D, Plebani M. Analytical performances of a chemiluminescence immunoassay for SARS-CoV-2 IgM/IgG and antibody kinetics. Clin Chem Lab Med 2020; (ahead of print). doi:10.1515/cclm-2020-0443

9. Zhao J, Yuan Q, Wang H, Liu W, Liao X, Su Y, et al. Antibody responses to SARS-CoV-2 in patients of novel coronavirus disease 2019. Clin Infect Dis. 2020; (ahead of print). doi:10.1093/cid/ciaa344.

10. Guo L, Ren L, Yang S, Xiao M, Chang, Yang F, et al. Profiling Early Humoral Response to Diagnose Novel Coronavirus Disease (COVID-19). Clin Infect Dis. 2020; (ahead of print). doi:10.1093/cid/ciaa310.

11. Sethuraman N, Jeremiah SS, Ryo A. Interpreting Diagnostic Tests for SARS-CoV-2 [published online ahead of print, 2020 May 6]. JAMA. 2020; (ahead of print). 10.1001/jama.2020.8259. doi:10.1001/jama.2020.8259

12. Lippi G, Salvagno GL, Pegoraro M, Militello V, Caloi C, Peretti A, et al. Assessment of immune response to SARS-CoV-2 with fully automated MAGLUMI 2019-nCoV IgG and IgM chemiluminescence immunoassays. Clin Chem Lab Med. 2020; (ahead of print). doi:10.1515/cclm-2020-0473.

13. Padoan A, Sciacovelli L, Basso D, Negrini D, Zuin S, Cosma C, et al. IgA-Ab response to spike glycoprotein of SARS-CoV-2 in patients with COVID-19: A longitudinal study. Clin Chim Acta. 2020;507:164–166. (ahead of printf) doi:10.1016/j.cca.2020.04.026.

14. Liu W, Liu L, Kou G, Zheng Y, Ding Y, Ni W et al. Evaluation of nucleocapside and spike protein-based ELISAs for detecting antibodies against SARS-CoV-2. J Clin Microbiolo 2020. 10.1128/JCM.00461-2.

15. To KK, Tsang OT, Leung WS, Tam AR, Wu TC, Lung DC, et al. Temporal profiles of viral load in posterior oropharyngeal saliva samples and serum antibody responses during infection by SARS-CoV-2: an observational cohort study. Lancet Infect Dis 2020;20:565–74.

16. Fagiuoli S, Lorini FL, Remuzzi G. Covid-19 Bergamo Hospital Crisis Unit.Adaptations and Lessons in the Province of Bergamo. N Engl J Med. 2020;382(21):e71 (ahead of print). doi:10.1056/NEJMc2011599.

17. Long QX, Liu BZ, Deng HJ, Wu GC, Deng K, Chen YK, et al. Antibody responses to SARS-CoV-2 in patients with COVID-19. Nat Med. 2020. (ahead of print). doi:10.1038/s41591-020-0897-1.

18. Brochot E, Demey B, Touze A, Belouzard S, Dubuisson J, Schmit JL, et al. Anti-Spike, anti-Nucleocapsid and neutralizing antibodies in SARS-CoV-2 hospitalized patients and asymptomatic carriers. MedRxiv, 2020. https://doi.org/10.1101/2020.05.12.20098236.

19. Grifoni A, Weiskopf D, Ramirez S., Mateus J., Dan J, Rydyznski C, et al. Targets of T cell responses to SARS-CoV-2 coronavirus in humans with COVID-19 disease and unexposed individuals. Cell. 2020; (ahead of print). https://doi.org/10.1016/j.cell.2020.05.015.

20. Braun J, Loyal L, Frentsch M, Wendisch D, Georg P, Kurth F, et al. Presence of SARS-CoV-2 reactive T cells in COVID-19 patients and healthy donors. https://doi.org/10.1101/2020.04.17.20061440

21. To KK, Tsang OT, Leung WS, Tam AR, Wu TC, Lung DC, et al. Temporal profiles of viral load in posterior oropharyngeal saliva samples and serum antibody responses during infection by SARS-CoV-2: an observational cohort study. Lancet Infect Dis 2020;20:565–74.

22. Jiang S, Hillyer C, Du L. Neutralizing Antibodies against SARS-CoV-2 and other Human Coronaviruses. Trends in Immunology 2020; 41: 355–359.

23. Plebani M, Padoan A, Negrini D, Carpinteri B, Sciacovelli L. Diagnostic performances and thresholds: the key to harmonization in serological SARS-CoV-2 assays? Clin Chim Acta, 2020 (accepted for publication).

